# A meta-analysis of complications of thread lifting

**DOI:** 10.1101/2025.03.10.25323087

**Authors:** Xiao-Cheng Zhou, Shu-Bo Zhuang

## Abstract

**Objective:** This study aims to systematically review and perform a meta-analysis on the complications associated with thread lifting, a popular minimally invasive aesthetic procedure, to determine the incidence and types of adverse effects.

**Methods:** A comprehensive literature search was conducted across major medical databases including PubMed, Embase, and Web of Science, covering all publications up to April 1, 2024. The search terms included “thread lift,” “suture lift,” “barbed suture,” “facelift,” and “nonsurgical facelift,” combined with “complication” or “adverse effect.” Only prospective or retrospective cohort studies, clinical randomized controlled trials (RCTs), and case series published in English were included. Studies were excluded if they were non-English, review articles, case reports, or conference abstracts with incomplete data. Data on authors, publication year, study design, sample size, patient demographics, follow-up duration, and complications were extracted and analyzed using R 4.2. Heterogeneity among studies was assessed using the I^2^ statistic.

**Results:** Our comprehensive search initially identified 537 articles. After removing duplicated records and rigorous title and abstract screening, 80 articles were further assessed, with 26 studies ultimately included in the final analysis, representing a collective sample of 2,827 patients. The meta-analysis highlighted varying incidence rates of complications associated with thread lifts: swelling was reported in 16% of cases, pain in 11%, skin dimpling or asymmetry pain in 7%, paresthesia in 10%, visible or palpable threads in 6%, infection in 2%, ecchymoses in 26% and thread exposure in 5%. The analysis also revealed high heterogeneity among the studies, with I^2^ values indicating substantial to high variability: swelling (I^2^ = 92%), skin dimpling (I^2^ = 76%), visible threads (I^2^ = 88%), and ecchymoses (I^2^ = 92%). Less common complications such as ear numbness and pinching sensation were reported in fewer studies, affecting approximately 5% and 7% of patients, respectively.

**Conclusion:** Thread lifting, while generally safe, does carry a risk of several complications, which vary widely in their occurrence. This meta-analysis provides a detailed overview of the risk profile of thread lifting procedures, highlighting the need for careful patient selection and technique mastery by practitioners. The findings underscore the importance of setting realistic patient expectations and preparing for potential adverse effects.

## introduction

Over the last several decades, cosmetic surgery has expanded significantly due to heightened societal emphasis on aesthetic appearance[1–3]. Within this burgeoning field, thread lifting has emerged as a pivotal technique, characterized by its minimal invasiveness and substantial efficacy in facial rejuvenation[4, 5]. This procedure involves the strategic placement of absorbable sutures beneath the skin to leverage the tension created by the threads to elevate and reposition the skin and subdermal structures, thereby delivering a pronounced tightening and lifting effect[6]. Notable for its operational simplicity, relatively brief recovery timeframe, minimal invasiveness, and immediate visible results, thread lifting has secured its role as a cornerstone in the repertoire of modern non-surgical aesthetic interventions[7, 8].

Despite its apparent benefits, the potential for adverse effects associated with thread lifting necessitates careful consideration. The literature documents various complications, including local infections[9], migration of the threads[10], skin dimpling[11–13], tissue fibrosis[14], appearance anomalies[15], and suture breakage[16, 17]. These issues not only compromise the aesthetic outcomes but may also necessitate further medical interventions, potentially imposing significant psychological and financial burdens on patients.

Given these considerations, our study conducts a comprehensive meta-analysis to rigorously quantify the incidence of complications associated with thread lifting. By systematically collating and analyzing data from a multitude of studies, we aim to furnish a robust scientific foundation that will aid clinicians in making informed procedural choices and enable patients to better understand the associated risks. This informed approach is intended to optimize patient outcomes and satisfaction by mitigating potential complications through enhanced procedural strategies and patient education.

## Methods

### literature search strategy

We conducted this systematic review in accordance with the Preferred Reporting Items for Systematic Reviews and Meta Analyses (PRISMA) guidelines, to ensure comprehensiveness and systematic coverage, this study’s literature search encompassed the following major medical databases: PubMed, Embase, and Web of Science. The search covered the period from the inception of each database until April 1, 2024. The search terms combined specific technique names and related medical terms, including “thread lift,” “suture lift,” “barbed suture,” “facelift,” “nonsurgical facelift,” as well as “complication” or “adverse effect.” To enhance the breadth of the search, both MeSH terms and free text were utilized to adapt to the search features of each database, in addition, we will manually check the references to discover potentially available studies. Detailed search strategies can be found in the appendix Table 1.

**Table 1.** Basic characteristics of included studies.

| Author | year | Study Design | n | Follow up | Mean age (range) |
| --- | --- | --- | --- | --- | --- |
| R. F. Abraham | 2009<sup>[18]</sup> | Retrospective | 33 | 21 m | NA |
| M. S. El-Mesidy | 2020<sup>[19]</sup> | Prospective | 20 | 2 m | 48(39-62) |
| M. Fukaya | 2017<sup>[20]</sup> | Retrospective | 100 | 12 m | 45 ( 25-64 ) |
| S. E. Han | 2016<sup>[21]</sup> | Prospective | 20 | 12 m | 43(32-61) |
| M. S. Kaminer | 2008<sup>[22]</sup> | Retrospective | 20 | 11.5 m | 58 ( 39-73 ) |
| S. H. Kang | 2017<sup>[23]</sup> | Retrospective | 39 | 6 m | 45(31-70) |
| H. H. Kwon | 2019<sup>[24]</sup> | Retrospective | 25 | 4 m | 33-67 |
| C. Le Louarn | 2018<sup>[25]</sup> | Retrospective | 342 | 13.6 | NA |
| H. Lee | 2018<sup>[26]</sup> | Retrospective | 35 | 12 m | NA |
| M. P. Ogilvie | 2018<sup>[27]</sup> | Prospective | 100 | 6 m | 62(41-87) |
| Y. J. Park | 2021<sup>[28]</sup> | Prospective | 73 | 24 m | 51(31-67) |
| J. D. Rachel | 2010<sup>[29]</sup> | Retrospective | 29 | 12 m | 54(32-68) |
| S. Rezaee Khiabanloo | 2020<sup>[30]</sup> | Prospective | 58 | 6 m | 53(30-76) |
| A. Santorelli | 2021<sup>[31]</sup> | Retrospective study | 60 | 9.8 m | 51(32-71) |
| S. Sarigul Guduk | 2018<sup>[32]</sup> | Retrospective | 148 | 22 m | 45(21-67) |
| A. Savoia | 2014<sup>[33]</sup> | Prospective | 37 | 24 m | 37-65 |
| D. H. Suh | 2015<sup>[34]</sup> | Retrospective | 31 | 24 w | 44 |
| M. A. Sulamanidze | 2002<sup>[35]</sup> | Prospective | 186 | 30 m | 21-77 |
| R. Wanitphakdeedecha | 2021<sup>[36]</sup> | Prospective | 25 | 12 m | 30-55 |
| K. Wattanakrai | 2020<sup>[37]</sup> | Prospective | 21 | 24 m | NA |
| N. Yu | 2020<sup>[38]</sup> | Retrospective | 46 | 12 m | 50 ( 38-64 ) |
| J. de Benito | 2011<sup>[39]</sup> | Prospective | 316 | 18 m | 47(28-66) |
| P. B. Garvey | 2009<sup>[40]</sup> | Retrospective | 72 | 24 m | 57 |
| S. Lee and N. Isse | 2005<sup>[41]</sup> | Retrospective | 44 | 9 m | NA |
| A. Z. E. D. Badin | 2005<sup>[42]</sup> | Prospective | 52 | 18 m | 14-81 |
| B. Lycka | 2004<sup>[43]</sup> | Prospective | 350 | 24 m | NA |

m: month; y: year; w: week; NA: Not available.

### Inclusion and exclusion criteria

The inclusion criteria are as follows: the study subjects are patients undergoing thread lifting; the study design include prospective or retrospective cohort studies, clinical randomized controlled trials (RCTs), and case series; the study outcomes are all postoperative complications, including swelling, pain in 11%, skin dimpling, paresthesia, visible or palpable threads, infection, ecchymoses and thread exposure. etc. Exclusion criteria include non English literature, review articles, case reports, conference abstracts, and studies with incomplete data.

### Study screening

The initial phase of the literature selection process involved a thorough screening of titles and abstracts to identify studies that potentially aligned with the specific focus of our research on thread lifting and associated complications. This step effectively eliminated any publications that did not directly address the core themes of our study. Following this initial screening, two independent researchers conducted a detailed full-text review of the remaining articles. This comprehensive evaluation was critical to accurately assess each study’s relevance and adherence to our stringent inclusion and exclusion criteria. It ensured that only the most pertinent studies were considered for our analysis, thereby maintaining the integrity and scientific rigor of our research. In instances where the two reviewers disagreed on the eligibility of a particular study, a third-party expert was consulted to arbitrate and provide a decisive judgment. This arbitration process was an essential measure to guarantee impartiality and consensus in the final study selection, ensuring that all included research met our high standards for quality and relevance.

### Data Extraction

For this systematic review, detailed data were meticulously extracted from each selected study based on pre-designed tables to construct a robust dataset essential for our analysis. The extracted information included the names of the authors and the year the study was published, providing context and allowing for assessment of the research’s recency and relevance. The type of study design was noted to evaluate the methodological quality and applicability of the findings. The sample size of each study was recorded to understand the scale and statistical power of the results. We also gathered detailed baseline characteristics of the study populations, including age and gender, which are crucial for assessing the generalizability of the findings to broader populations. The duration of follow-up was extracted to determine the longitudinal reliability of the results and to assess the long-term implications of thread lifting complications.

Furthermore, the specific types of complications reported in the studies and their respective incidence rates were carefully documented. This information is vital for understanding the risk profile associated with thread lifting procedures and for informing clinical practice.The data extraction process was carried out independently by two researchers to minimize bias and ensure data integrity. Any discrepancies encountered during the data extraction phase were resolved through direct discussion between the researchers, facilitating a consensus-driven approach to ensure the accuracy and reliability of the data used in our analysis. This meticulous process underpins the validity of our research conclusions, contributing significantly to the existing body of knowledge on cosmetic surgery complications.

### quality assessment

The quality appraisal of the literature was based on a specific checklist, the “Quality Appraisal of Case Series Studies Checklist,” which originally consists of 20 items. In your study, 10 relevant questions from this checklist were selected for evaluation. Based on these 10 selected questions, the assessment criteria are as follows: High quality: If all 10 questions are answered affirmatively, the study is defined as high quality. Moderate quality: If eight or more of the questions are answered affirmatively, the study is defined as moderate quality. Low quality: If fewer than eight questions are answered affirmatively, the study is defined as low quality. In the event of any dispute regarding the eligibility or quality of the literature, a third party will adjudicate.

### Statistical Analysis

We extracted all outcomes involved in the studies included in our research, conducting meta-analyses only on those outcomes reported in three or more articles. The meta-analyses were executed using R 4.2 software, a robust tool for handling complex statistical data in systematic reviews. For assessing the incidence of complications, we used a random-effects model, which is particularly suitable given the expected variability across studies in terms of procedures, patient groups, and study settings.

To address and quantify heterogeneity, which reflects the degree to which the effect sizes vary among included studies, we employed the I^2^ statistic. This measure helps determine the suitability of pooling the individual studies’ results and interpreting the meta-analysis’s overall outcome. The heterogeneity was categorized into four levels: I^2^ < 25% was considered to reflect low heterogeneity; 25% ≤ I^2^ < 50% was indicative of moderate heterogeneity; 50% ≤ I^2^ < 75% suggested substantial heterogeneity; and I^2^ ≥ 75% was associated with high heterogeneity.

Statistical significance was determined at a threshold of P < 0.05. This conventional cut-off provides a balance between identifying genuine effects and minimizing the risk of false-positive results. Furthermore, to investigate potential publication bias, which could skew the meta-analysis results, we utilized funnel plots. These plots are particularly effective in identifying asymmetries that could indicate bias. However, to increase the reliability of these tests, we limited funnel plot analyses to outcomes reported in ten or more studies, ensuring a robust sample size for detecting genuine trends in data dispersion. This meticulous approach to data analysis enhances the validity and credibility of our findings, providing a comprehensive understanding of the risk associated with thread lifting procedures.

## Results

### Study selection results

In the preliminary search, approximately 537 related articles were retrieved. After removing 65 duplicate articles, we filtered out an additional 392 articles based on their titles and abstracts. The remaining 80 articles were further screened through full-text reviews, and 54 articles were excluded mainly because they were reviews, letters, abstract literature, unrelated to the research topic, or had no relevant outcome variables. Finally, 26 studies were included for meta-analysis. Figure 1 shows the process of screening literature.

**Figure 1.**
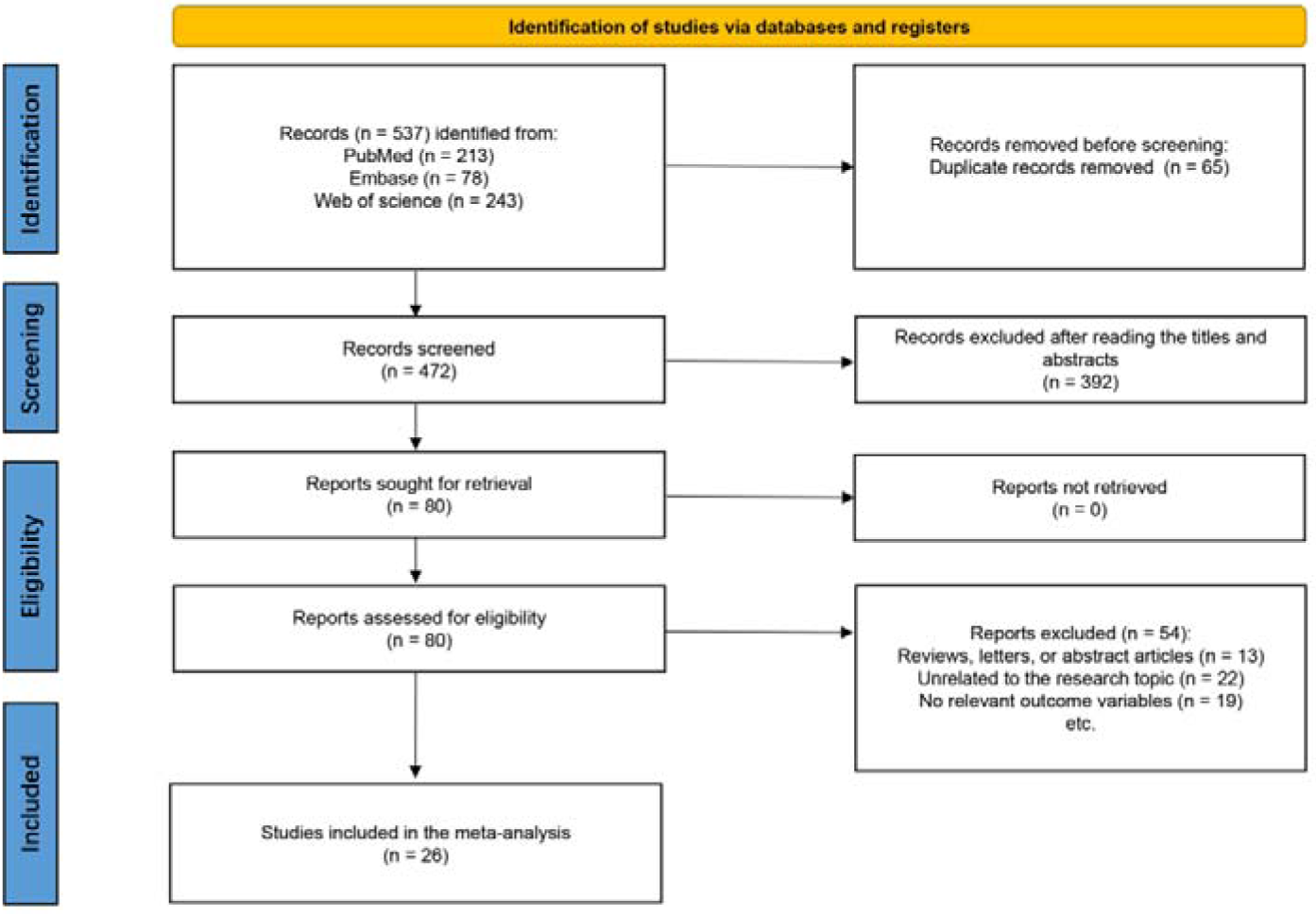
Literature screening process diagram

### Characteristics of Studies

This meta-analysis encompassed data from a total of 29 studies, comprising 2,827 patients across various investigations on thread lift procedures (Table 1). The studies ranged in design, including retrospective, prospective studies, and comparative analyses, conducted between 2002 and 2021. The patient cohort exhibited a wide age range, from 14 to 87 years, reflecting a diverse demographic. Follow-up duration varied from 4 weeks to 30 months, providing a comprehensive overview of complications associated with thread lift procedures across different time frames.

### Incidences of Complications

Figure 2 illustrates the occurrence of swelling following thread lift procedures. Among the 26 studies included in our analysis, 15 reported swelling. Based on a random-effects model, the overall proportion of swelling following facial lift surgery is 0.16 (95% CI: 0.7 to 0.34). However, substantial heterogeneity existed among the studies (I^2^=92%).

**Figure 2.**
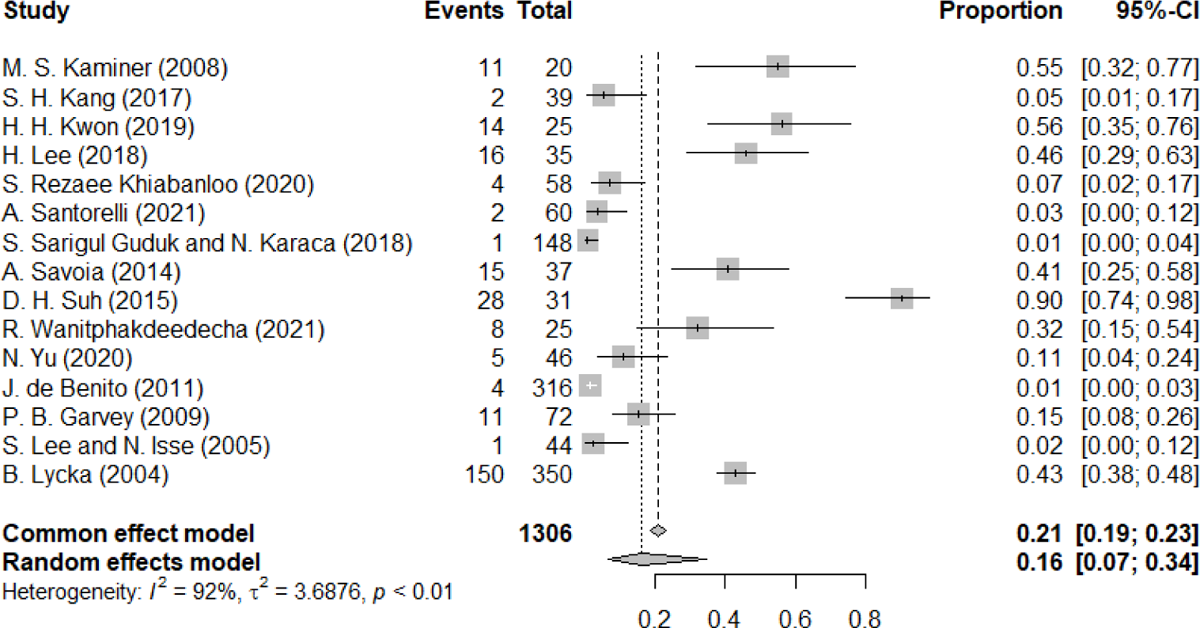
The forest plot of swelling

Figure 3 shows the occurrence of pain after thread lifting. A total of 6 studies reported this complication. This forest plot for the complication of pain following facial lifting shows a pooled proportion of 0.11 (95% CI: 0.04 to 0.24) using a random effects model, with significant heterogeneity (I^2^ = 86%).

**Figure 3.**
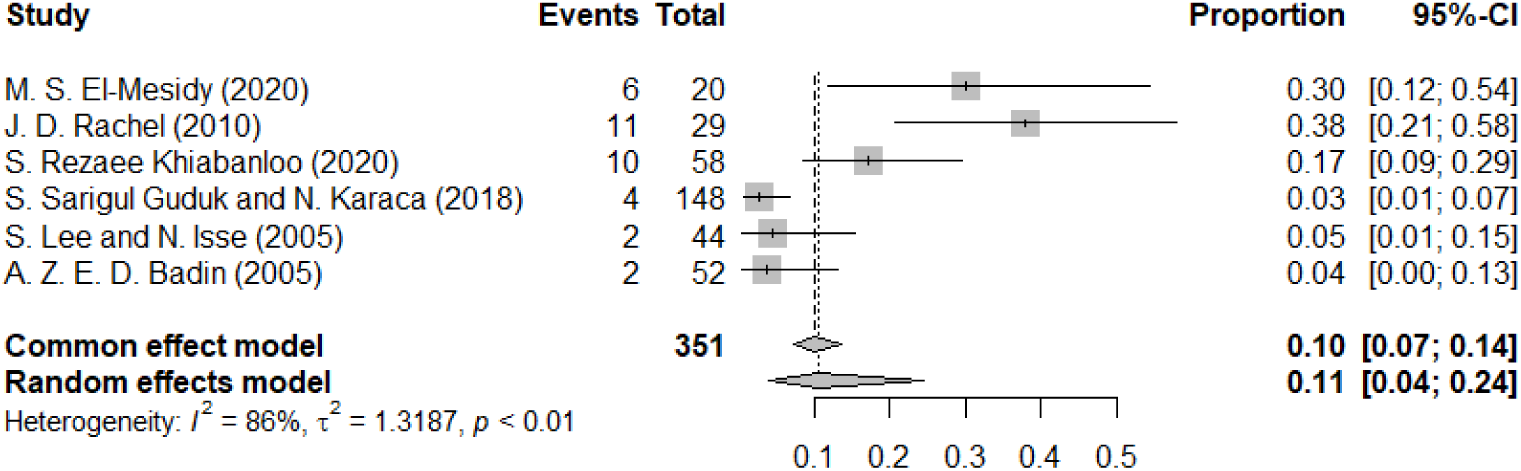
The forest plot of pain

Skin dimpling or asymmetry is a common complication of thread lifting. 16 studies reported this adverse event, mainly skin dimpling. The forest plot depicts the incidence of skin dimpling or asymmetry following facial lifting procedures across multiple studies (Figure 4). The random effects model estimated a pooled proportion of 0.07 (95% CI: 0.04 to 0.12), suggesting that approximately 7% of patients experience postoperative skin dimpling or asymmetry. Heterogeneity among the studies was observed (I^2^ = 76%), indicating variability in outcomes across different research settings.

**Figure 4.**
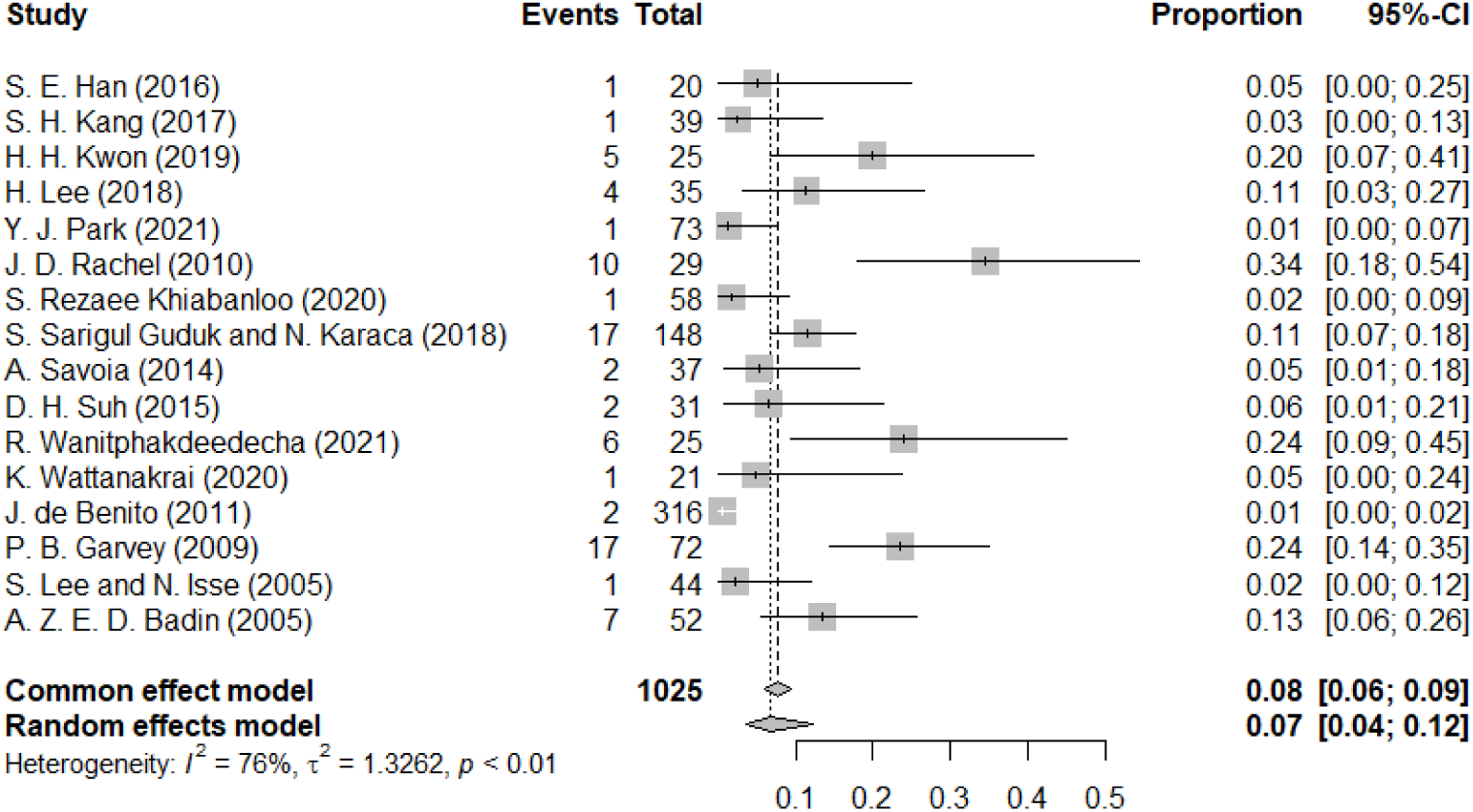
The forest plot of skin dimpling or asymmetry pain

Figure 5 is the forest map of paresthesia. The forest plot indicates the results for the complication of paresthesia following facial lifting procedures across a selection of studies. The random effects model estimates a pooled proportion of 0.06 (95% CI: 0.01 to 0.26), signifying a moderate average incidence rate with substantial variability, as reflected by the high heterogeneity (I^2^ = 87%).

**Figure 5.**
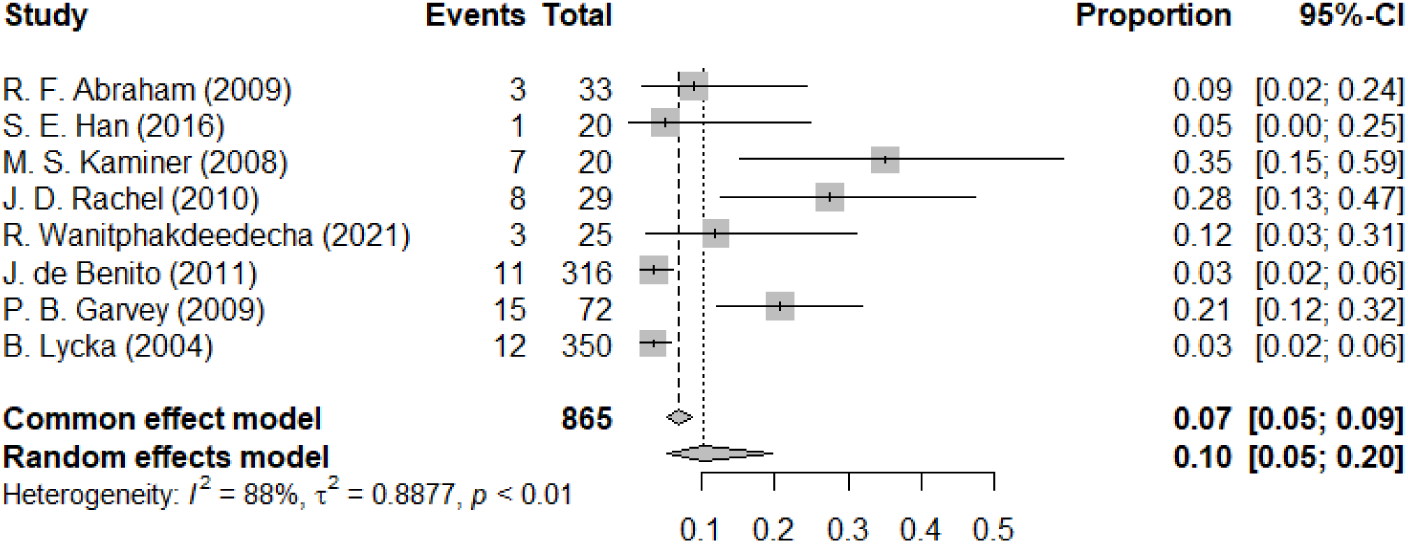
The forest plot of paresthesia

Figure 6 depicts another common complication. The forest plot for the complication of visible or palpable threads, also described as a visible knot, following facial lifting procedures demonstrates a random effects model pooled proportion of 0.10 (95% CI: 0.05 to 0.20), indicating that an average of 10% of patients might experience this issue post-procedure. The common effect model suggests a slightly lower pooled proportion of 0.07 (95% CI: 0.05 to 0.09). The heterogeneity among the studies is substantial (I^2^ = 88%), suggesting considerable variation in the reported rates of this complication across different studies.

**Figure 6.**
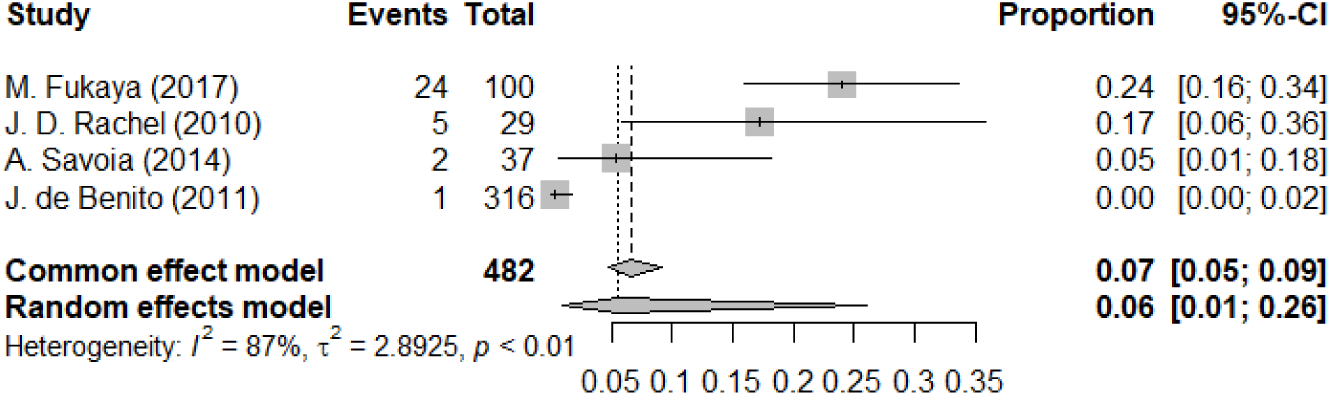
The forest plot of visible or palpable threads/visible knots

Figure 7 is a meta-analysis plot of infection. Only 3 studies reported infection. The overall incidence rate was low, with a pooled effect size of 0.02 (0.01-0.06). Figure 8 depicts this complication of ecchymoses. The forest plot for the incidence of ecchymoses as a postoperative complication shows individual study proportions ranging broadly with a pooled estimate of 0.26 (95% CI: 0.11 to 0.51) under a random effects model, reflecting considerable variation in outcomes, as indicated by a high I^2^ value of 92%.

**Figure 7.**
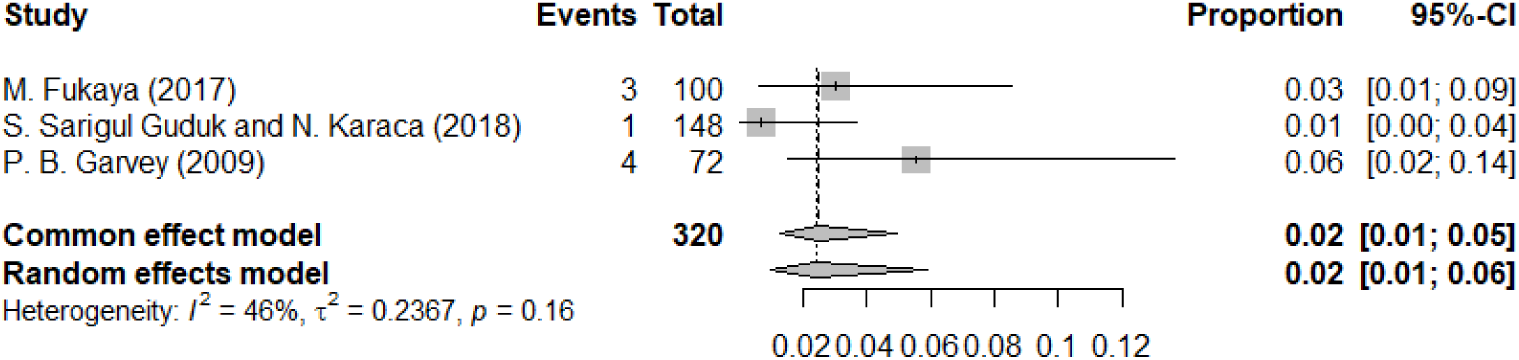
The forest plot of infection

**Figure 8.**
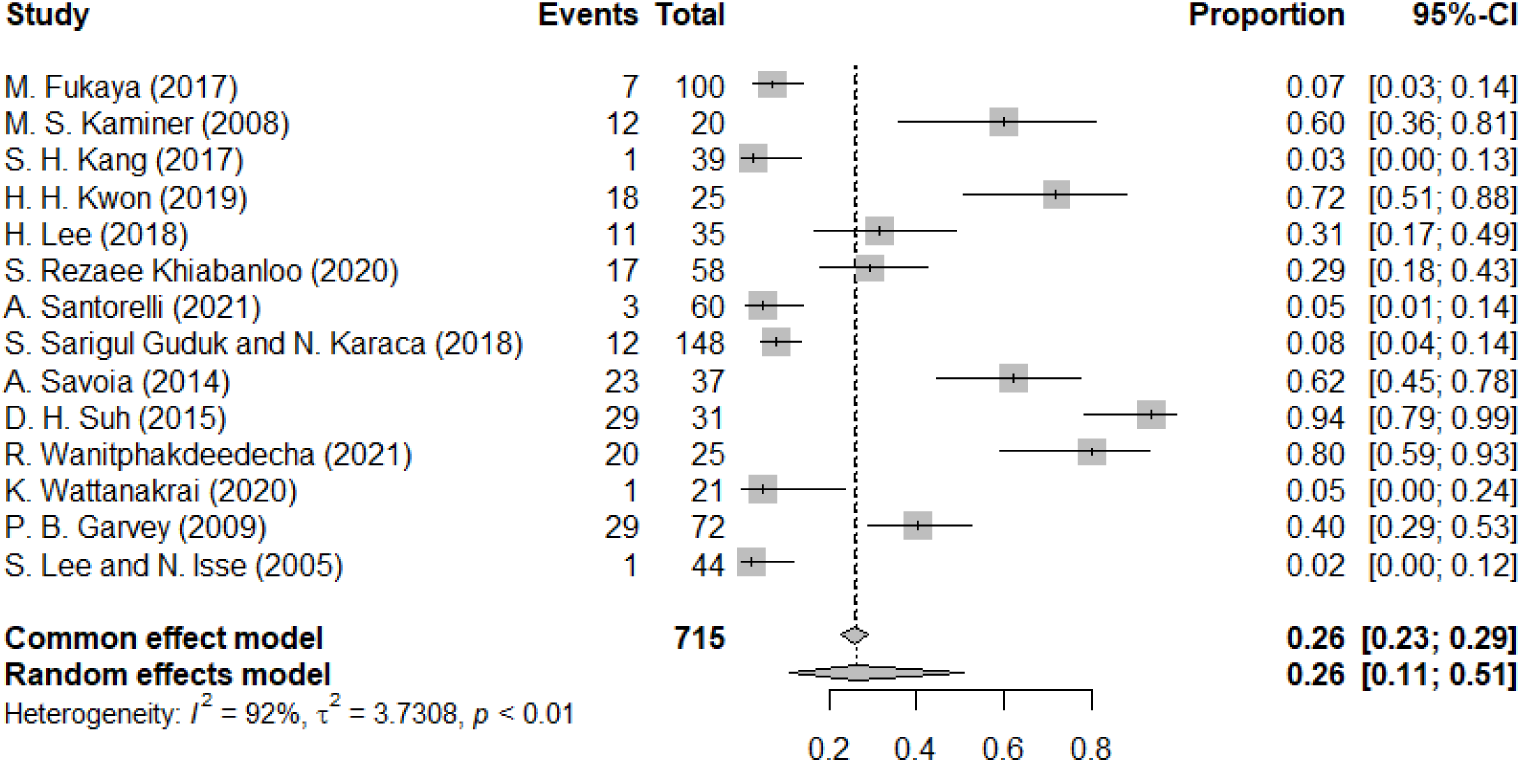
The forest plot of ecchymoses

Figure 9 illustrates this complication of thread exposure. The forest plot summarizes the occurrence of thread exposure as a complication post-facial lifting procedures, showing an estimated pooled proportion of 0.05 (95% CI: 0.03 to 0.08) in the random effects model and 0.05 (95% CI: 0.03 to 0.07) in the common effect model. The heterogeneity across the included studies is moderate (I^2^ = 44%), with individual study effects varying but generally indicating a low incidence of this particular complication.

**Figure 9.**
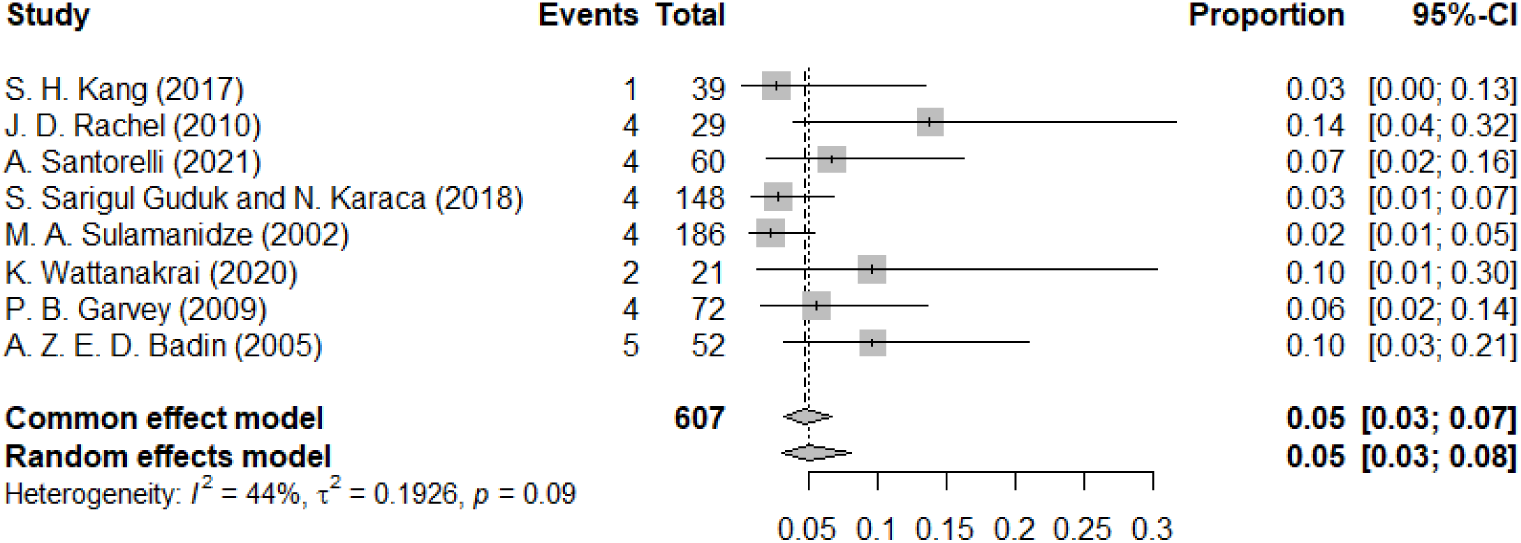
The forest plot of thread exposure

Other less common complications include ear numbness, Pinching sensation, Malar eminence accentuation, small hemorrhage is illustrated in Table 2.

**Table 2.**
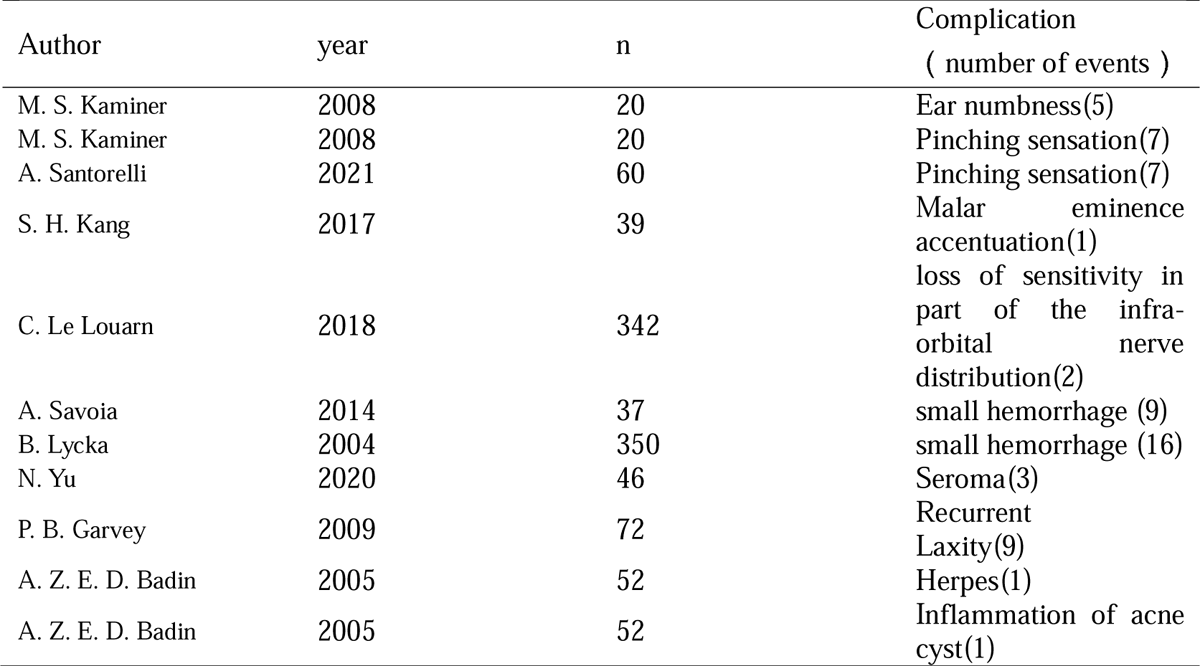
Other complications.

### Publication bias assessment and literature quality assessment

Figure 10 shows the funnel plot of swelling, skin dimpling, and ecchymosis. The graph is not completely symmetrical and has a large bias. Table 3 shows the literature quality evaluation results. The overall research quality is relatively high.

**Figure 10.**
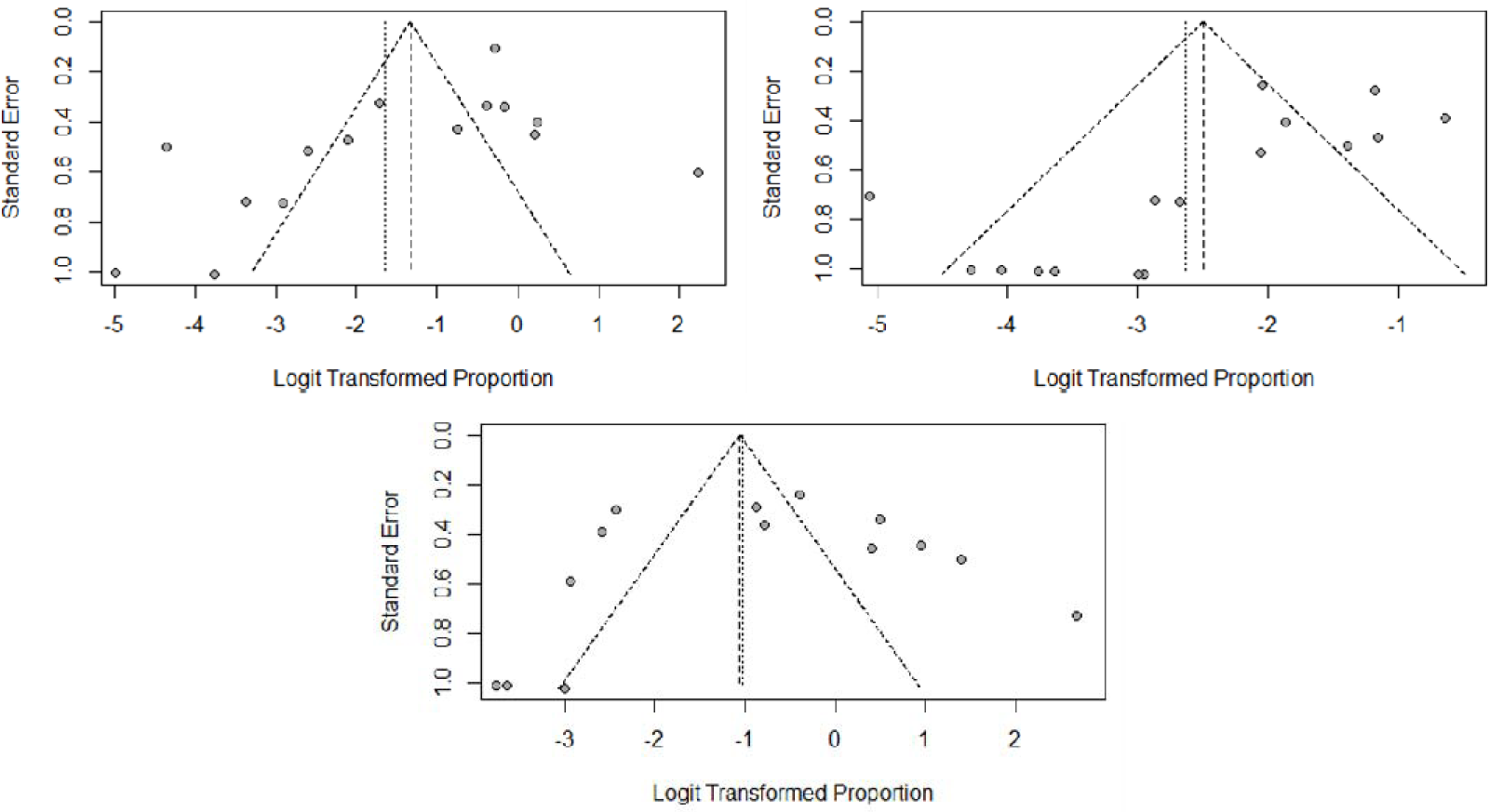
Funnel plot of swelling, skin dimpling, and ecchymoses

**Table 3.**
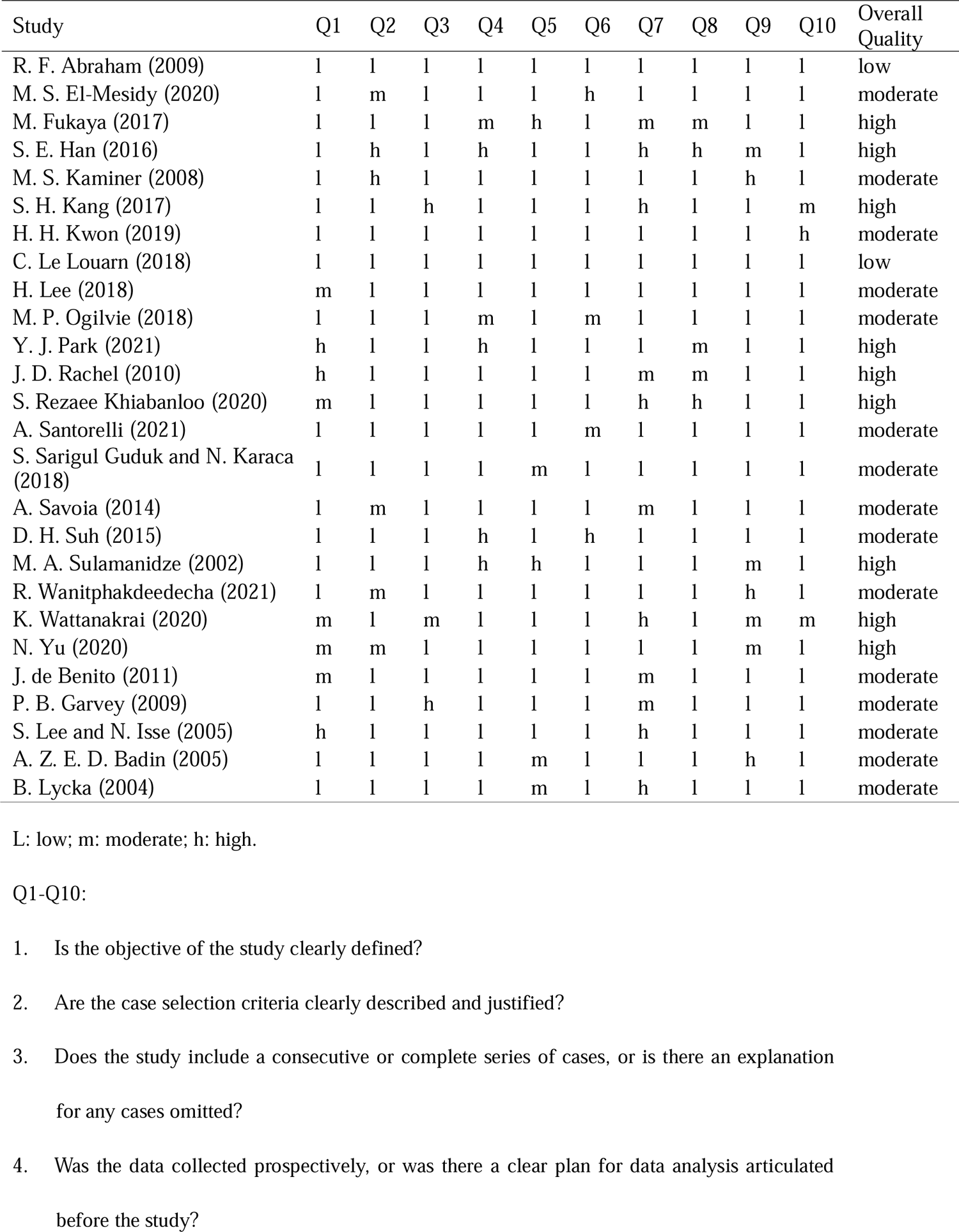
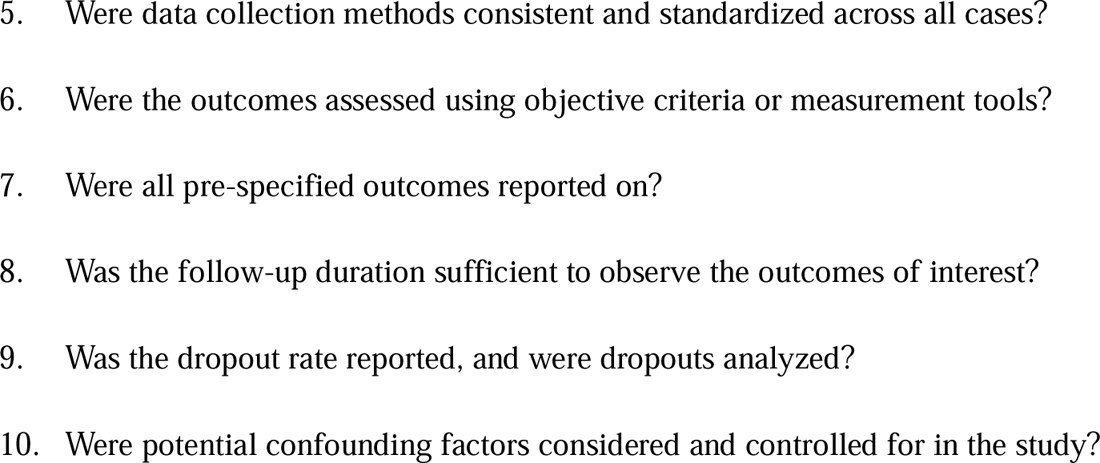
Literature quality evaluation.

## Discussion

The impetus for conducting this study stemmed from a growing interest in the aesthetic community regarding the safety and efficacy of thread lifting, a procedure that has gained rapid popularity due to its minimally invasive nature and immediate results. This meta-analysis was initiated to systematically assess the range and frequency of complications associated with thread lifting, a subject of increasing concern amongst both practitioners and patients. Our findings have elucidated that while thread lifting is generally a safe procedure, complications such as swelling, skin dimpling, visible threads, and ecchymoses occur at variable rates. Notably, the incidence rate for swelling was found to be the highest, reported at 34%, followed by ecchymoses at 26%. These results underscore the necessity for thorough pre-procedural planning and patient counseling to mitigate these risks effectively.

Our study corroborates several findings from previous research, primarily confirming that thread lifting, while effective, is not devoid of risks[44–46]. Similar studies in the past have highlighted complications like minor swelling[39, 43] and bruising[34, 40] as common immediate postoperative occurrences, which aligns with our observations of high incidence rates for these issues. Additionally, the complications related to thread visibility and migration reported in earlier studies were also consistently observed in our analysis, further validating the persistent nature of these complications across different study cohorts and procedural techniques.

Despite its strengths, our study has several limitations that warrant mention. First, the inherent heterogeneity of the included studies, in terms of both procedural techniques and patient demographics, may have influenced the complication rates observed. The variability in thread types used, practitioner skill, and patient skin types across studies could have contributed to the differences in complication incidences. Moreover, our analysis was confined to studies published in English, potentially omitting relevant data from non-English publications that could provide additional insights into the complications associated with thread lifting. Finally, the retrospective nature of many included studies may introduce recall bias, particularly in self-reported outcomes such as pain or minor complications, which might not always be accurately documented.

Given the findings and the limitations noted, it is imperative for practitioners to approach thread lifting with a critical eye towards optimizing techniques and selecting candidates. Practitioners should be well-versed in the various suture materials and techniques to tailor their approach to the individual patient’s anatomy and desired outcomes, potentially reducing the incidence of complications. Patient selection should also involve a thorough assessment of skin quality and a detailed discussion about realistic expectations and possible adverse outcomes. Further research, particularly prospective studies with standardized protocols and longer follow-up periods, will be crucial in advancing our understanding of the long-term safety and efficacy of thread lifting.

## Conclusion

This meta-analysis has highlighted the complications associated with thread lifting, underscoring its effectiveness as a minimally invasive aesthetic procedure alongside its potential risks. Swelling, ecchymoses, visible threads, and skin dimpling emerged as common complications, indicating the need for meticulous technique and patient selection to mitigate these issues. Our findings stress the importance of thorough pre-procedural consultations and realistic discussions with patients about the risks involved. By improving practitioner skills and patient understanding, the safety and satisfaction outcomes of thread lifting can be enhanced. Future research should focus on standardizing methods and broadening patient demographics to further understand and refine the procedure’s safety profile.

## Data Availability

All data produced in the present work are contained in the manuscript

---

**Appendix Table 1.** Search strategies in Pubmed, Embase and Web of Science.

| Search number | Queries |
| --- | --- |
| Queries in PubMed |  |
| #1 | "thread lift" [Mesh] OR "suture lift" [Title/Abstract] OR "barbed suture" [Title/Abstract]<br>OR "facelift" [Title/Abstract] OR "nonsurgical facelift" [Title/Abstract]) |
| #2 | "complication" [Title/Abstract] OR "adverse effect" [Title/Abstract]) |
| #3 | #1 AND #2 |
| Queries in Embase |  |
| #1 | 'thread lift'/exp OR 'suture lift': ti, ab OR 'barbed suture': ti, ab OR 'facelift': ti, ab OR<br>'nonsurgical facelift': ti, ab |
| #2 | 'complication': ti, ab OR 'adverse effect': ti,ab |
| #3 | #1 AND #2 |
| Queries in WOS |  |
| #1 | (((((TS = (thread lift)) OR TS = (suture lift)) OR TS = (barbed suture)) OR TS = (facelift))<br>OR TS = (nonsurgical facelift)) |
| #2 | ((TS = (cognitive impairment)) OR TS = (cognitive dysfunction)) |
| #3 | #1 AND #2 |

## References

[1] Rohrich R J. Plastic Surgery: Staying inside the Lines [J]. Plast Reconstr Surg, 2021, 148(5S): 18S–19S.

[2] Wu H, Wang G, Shang Y, et al. Organoids and Their Research Progress in Plastic and Reconstructive Surgery [J]. Aesthetic Plast Surg, 2023, 47(2): 880–891.

[3] Bailey C M, Goldman J J. Discussion: Health Care Value in Plastic Surgery Practice [J]. Plast Reconstr Surg, 2024, 153(5): 1184–1185.

[4] Hong G W, Kim S B, Park S Y, et al. Basic concepts in facial and neck thread lifting procedures [J]. Skin Res Technol, 2024, 30(4): e13673.

[5] Park J H, Jeong J W, Park J U. Advanced Facial Rejuvenation: Synergistic Effects of Lower Blepharoplasty and Ultrasound Guided Mid-Face Lift Using Polydioxanone (PDO) Threads [J]. Aesthetic Plast Surg, 2024.

[6] Hong G W, Kim S B, Park S Y, et al. Why do marionette lines appear? Exploring the anatomical perspectives and role of thread-based interventions [J]. Skin Res Technol, 2024, 30(4): e13676.

[7] Yi K H. What are filling (volumizing) threads? [J]. Skin Res Technol, 2024, 30(3): e13658.

[8] Li Z, Wu H, Yang Z, et al. Combining Liposuction and Thread-Lifting for Middle-Lower Facial Rejuvenation [J]. Aesthetic Plast Surg, 2024.

[9] Kim H J, Lee S J, Lee J H, et al. Clinical Features of Skin Infection After Rhinoplasty with Only Absorbable Thread (Polydioxanone) in Oriental Traditional Medicine: A Case Series Study [J]. Aesthetic Plast Surg, 2020, 44(1): 139–147.

[10] Kasai H, Yashiro K, Kawahara Y. Multiple ulcers on the face due to infection after thread-lifting [J]. J Dermatol, 2018, 45(12): e336–e337.

[11] Oh C H, Jang S B, Kang C M, et al. Buttock Lifting Using Elastic Thread (Elasticum(®)) with a New Classification of Gluteal Ptosis [J]. Aesthetic Plast Surg, 2018, 42(4): 1050–1058.

[12] Li Y L, Li Z H, Chen X Y, et al. Facial Thread Lifting Complications in China: Analysis and Treatment [J]. Plast Reconstr Surg Glob Open, 2021, 9(9): e3820.

[13] Sahan A, Karaosmanoglu N, Ozdemir Cetinkaya P. Is it possible to obtain long-lasting results with thread lift in the brow region? Introduction of a new suspension technique and evaluation of 50 patients [J]. J Cosmet Dermatol, 2023, 22(6): 1863–1869.

[14] Navarro-Viana F. Rhytidectomy assisted with ultrasound techniques: the ultra-lipo-lift technique [J]. Aesthetic Plast Surg, 2001, 25(3): 175–180.

[15] De Benito J, Pizzamiglio R, Theodorou D, et al. Facial rejuvenation and improvement of malar projection using sutures with absorbable cones: surgical technique and case series [J]. Aesthetic Plastic Surgery, 2011, 35(2): 248–253.

[16] Sapountzis S, Kim J H, Li T S, et al. Successful treatment of thread-lifting complication from APTOS sutures using a simple MACS lift and fat grafting [J]. Aesthetic Plast Surg, 2012, 36(6): 1307–1310.

[17] De Clermont-Tonnerre E, Guinier C, Klap B, et al. Awareness of Facial Thread-Lifting: Report of a Rare Case [J]. Aesthet Surg J, 2022, 42(5): Np363–np364.

[18] Abraham R F, Defatta R J, Williams E F, 3rd. Thread-lift for facial rejuvenation: assessment of long-term results [J]. Arch Facial Plast Surg, 2009, 11(3): 178–183.

[19] El-Mesidy M S, Alaklouk W T, Azzam O A. Nasolabial fold correction through cheek volume loss restoration versus thread lifting: a comparative study [J]. Arch Dermatol Res, 2020, 312(7): 473–480.

[20] Fukaya M. Long-term effect of the insoluble thread-lifting technique [J]. Clin Cosmet Investig Dermatol, 2017, 10: 483–491.

[21] Han S E, Go J Y, Pyon J K, et al. A Prospective evaluation of outcomes for midface rejuvenation with mesh suspension thread: “REEBORN lift” [J]. J Cosmet Dermatol, 2016, 15(3): 254–259.

[22] Kaminer M S, Bogart M, Choi C, et al. Long-term efficacy of anchored barbed sutures in the face and neck [J]. Dermatologic Surgery: Official Publication For American Society For Dermatologic Surgery [et Al], 2008, 34(8): 1041–1047.

[23] Kang S H, Byun E J, Kim H S. Vertical Lifting: A New Optimal Thread Lifting Technique for Asians [J]. Dermatol Surg, 2017, 43(10): 1263–1270.

[24] Kwon H H, Choi S C, Park G H, et al. Clinical Evaluations of a Novel Thread Lifting Regimen Using Barbed Polyglyconate Suture for Facial Rejuvenation: Analysis Using a 3-Dimensional Imaging System [J]. Dermatol Surg, 2019, 45(3): 431–437.

[25] Le Louarn C. Concentric Malar Lift in the Management of Lower Eyelid Rejuvenation or Retraction: A Clinical Retrospective Study on 342 Cases, 13 Years After the First Publication [J]. Aesthetic Plast Surg, 2018, 42(3): 725–742.

[26] Lee H, Yoon K, Lee M. Outcome of facial rejuvenation with polydioxanone thread for Asians [J]. J Cosmet Laser Ther, 2018, 20(3): 189–192.

[27] Ogilvie M P, Few J W, Jr., Tomur S S, et al. Rejuvenating the Face: An Analysis of 100 Absorbable Suture Suspension Patients [J]. Aesthet Surg J, 2018, 38(6): 654–663.

[28] Park Y J, Cha J H, Han S E. Maximizing Thread Usage for Facial Rejuvenation: A Preliminary Patient Study [J]. Aesthetic Plast Surg, 2021, 45(2): 528–535.

[29] Rachel J D, Lack E B, Larson B. Incidence of complications and early recurrence in 29 patients after facial rejuvenation with barbed suture lifting [J]. Dermatol Surg, 2010, 36(3): 348–354.

[30] Rezaee Khiabanloo S, Nabie R, Aalipour E. Outcomes in thread lift for face, neck, and nose; A prospective chart review study with APTOS [J]. J Cosmet Dermatol, 2020, 19(11): 2867–2876.

[31] Santorelli A, Cerullo F, Cirillo P, et al. Mid-face reshaping using threads with bidirectional convergent barbs: A retrospective study [J]. J Cosmet Dermatol, 2021, 20(6): 1591–1597.

[32] Sarigul Guduk S, Karaca N. Safety and complications of absorbable threads made of poly-L-lactic acid and poly lactide/glycolide: Experience with 148 consecutive patients [J]. J Cosmet Dermatol, 2018, 17(6): 1189–1193.

[33] Savoia A, Accardo C, Vannini F, et al. Outcomes in thread lift for facial rejuvenation: a study performed with happy lift™ revitalizing [J]. Dermatology and Therapy, 2014, 4(1): 103–114.

[34] Suh D H, Jang H W, Lee S J, et al. Outcomes of polydioxanone knotless thread lifting for facial rejuvenation [J]. Dermatol Surg, 2015, 41(6): 720–725.

[35] Sulamanidze M A, Fournier P F, Paikidze T G, et al. Removal of facial soft tissue ptosis with special threads [J]. Dermatol Surg, 2002, 28(5): 367–371.

[36] Wanitphakdeedecha R, Yan C, Ng J N C, et al. Absorbable Barbed Threads for Lower Facial Soft-Tissue Repositioning in Asians [J]. Dermatol Ther (Heidelb), 2021, 11(4): 1395–1408.

[37] Wattanakrai K, Chiemchaisri N, Wattanakrai P. Mesh Suspension Thread for Facial Rejuvenation [J]. Aesthetic Plast Surg, 2020, 44(3): 766–774.

[38] Yu N, Yu P, Liu Z, et al. Elastic thread modified minimal access cranial suspension lift for lower and middle third facial rejuvenation [J]. Medicine (Baltimore), 2020, 99(13): e19381.

[39] Sulamanidze M, Sulamanidze G, Vozdvizhensky I, et al. Avoiding complications with Aptos sutures [J]. Aesthetic Surgery Journal, 2011, 31(8): 863–873.

[40] Garvey P B, Ricciardelli E J, Gampper T. Outcomes in threadlift for facial rejuvenation [J]. Ann Plast Surg, 2009, 62(5): 482–485.

[41] Lee S, Isse N. Barbed polypropylene sutures for midface elevation: early results [J]. Arch Facial Plast Surg, 2005, 7(1): 55–61.

[42] Badin A Z E D, Forte M R C, Esilva O L. Scarless mid- and lower face lift [J]. Aesthetic Surgery Journal, 2005, 25(4): 340–347.

[43] Lycka B, Bazan C, Poletti E, et al. The emerging technique of the antiptosis subdermal suspension thread [J]. Dermatol Surg, 2004, 30(1): 41–44; discussion 44.

[44] Liao Z F, Yang W, Li X, et al. Infraorbital Rejuvenation Combined with Thread-Lifting and Non-cross-linked Hyaluronic Acid Injection: A Retrospective, Case-Series Study [J]. Aesthetic Plast Surg, 2023.

[45] Kim D M, Baek S W, Park J M, et al. Multifunctional PDO Thread Coated with Mg(OH)(2)/ZnO Nanoparticles and Asiaticoside for Improved Facial Lifting [J]. Pharmaceutics, 2023, 15(9).

[46] Goel A, Rai K. Non-surgical facelift-by PDO threads and dermal filler: A case report [J]. J Cosmet Dermatol, 2022, 21(10): 4241–4244.

